# Evaluation of eleven commercially available PCR kits for the detection of Monkeypox virus DNA

**DOI:** 10.1101/2022.10.14.22281096

**Authors:** Janine Michel, Angelina Targosz, Thomas Rinner, Daniel Bourquain, Annika Brinkmann, Jilian A. Sacks, Lars Schaade, Andreas Nitsche

**Author notes:** Correspondence : Janine Michel.

## Abstract

Prior to the international spread of Monkeypox in May 2022, PCR kits for the detection of Orthopoxviruses, and specifically for monkeypox virus, were rarely available. Here we describe the evaluation of eleven recently-developed commercially available PCR kits for the detection of Monkeypox virus DNA.

---

For roughly 50 years, human Monkeypox has been rarely detected outside of Central and Western Africa [1], with one larger outbreak in the US in 2003 linked to rodents imported from Ghana [2]. Individual cases have been reported in non-endemic regions, such as the UK, the US, Singapore and Israel, with links to travel to endemic countries, but with limited onward transmissions. Since May 2022, there has been an increasing number of cases of human Monkeypox worldwide, particularly in Europe and the US; this outbreak was declared a public health emergency of international concern (PHEIC) on 23 July 2022 [3].

Unfortunately, but often the case for rare and neglected diseases, there is a lack of quality-assured tools to combat Monkeypox disease, including reliable, commercially-available, test kits for diagnosis. Until recently, few PCR kits were available, many of which were designed to detect Orthopoxviruses (OPV) and/or Variola virus, the causative agent of smallpox. Hence, specialized laboratories historically rely on well-validated, in-house developed protocols for diagnosis.

The increased spread of human Monkeypox has triggered the development of PCR kits designed to detect either OPV – the genus of Monkeypox and other viruses such as VACV, and CPXV, zoonotic viruses that can cause sporadic human infections or self-limiting outbreaks, but also Variola virus – or to specifically detect MPXV. Since MPXV clinical samples have been rare in the past, kits are often validated by in silico comparison to published sequences.

To evaluate ready-to-use kits for MPXV diagnosis, an 18-specimen panel was established (table 1) and characterized using the diagnostic workflow of the German Consultant Lab for poxviruses (table 2, [4,5]). The panel included DNA from MPXV clade I, clade IIa and clade IIb, other OPV and VZV. All samples were analyzed in duplicate.

**Table 1:** Composition and preparation of the evaluation panel for PCR kits developed to detect Orthopoxviruses or Monkeypox virus.

| CT aim | Clade II 2012 [5] | Clade IIb 2022<sup>a</sup> | Clade I<sup>b</sup> | others |
| --- | --- | --- | --- | --- |
| 17 |  |  |  | VACV<sup>c</sup>, VZV<sup>d</sup> |
| 19 |  |  |  | CPXV<sup>e</sup> |
| 28 | X | X |  |  |
| 29 |  |  | X |  |
| 30 | X | X |  |  |
| 32 | X | X | X |  |
| 34 | X | X |  |  |
| 35 |  |  | X |  |
| 36 | X | X |  |  |
| 38 | X | X |  |  |

<sup>a</sup> 2022, MPXV/Germany/2022/ON/RKI305, OP494258. <sup>b</sup> MSF6, kindly provided by Hermann Meyer, IMB, Munich. <sup>c,e</sup> VACV Vaccinia Virus, CPXV Cowpoxvirus, CT determined with the rpo18 PCR [4]. <sup>d</sup> VZV Varicella Zoster Virus, CT determined by in-house PCR.

**Table 2:**
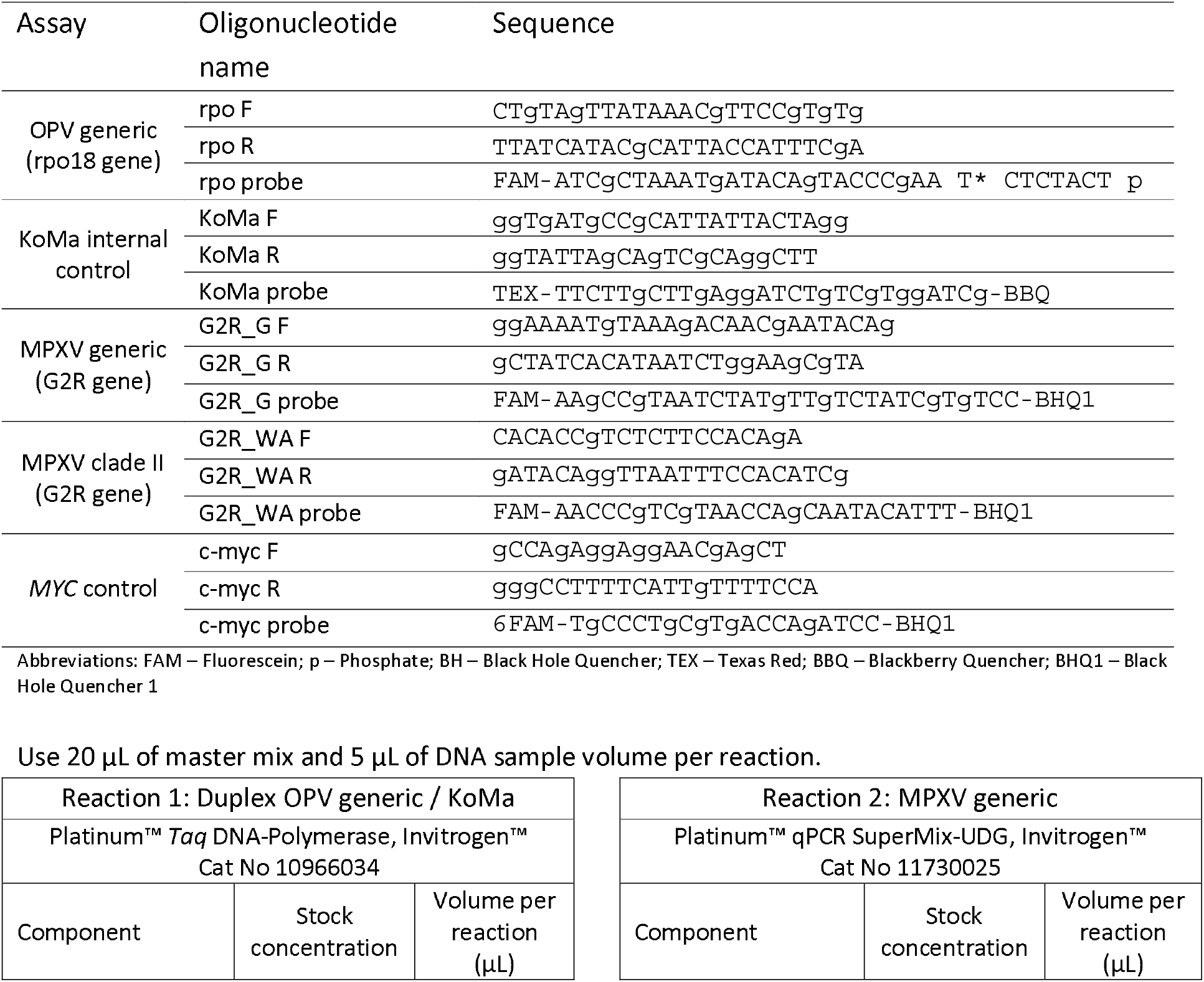
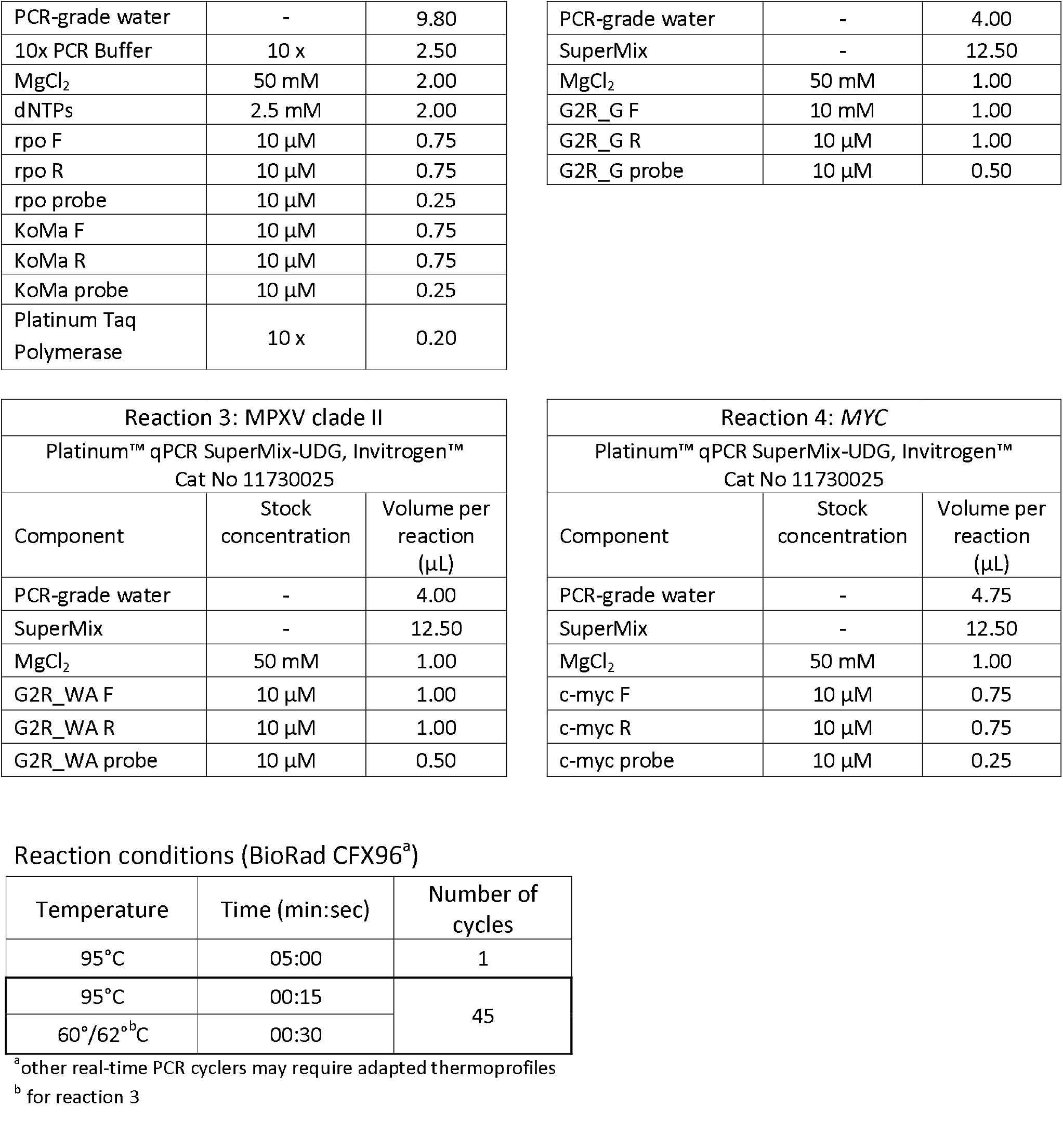
In-house protocol used as reference for detection of Monkeypox virus DNA

In total, eleven kits (A to L) were compared to the reference diagnostic workflow (table 2), which includes one generic OPV PCR (5), one MPXV-specific PCR, and one MPXV clade II-specific PCR [4]. Additionally, an inhibition control was spiked into the specimens prior to DNA extraction [5] and proper sampling was verified using a human DNA-specific PCR reaction [6]. All kits were used according to the manufacturer’s manual and the threshold was set to obtain the lowest possible CT value. For better comparability, all kits were run on the BioRad CFX 96 real-time cycler, which is compatible with the fluorophores used by all included tests, even if it was not specifically noted in the manual (table 3).

**Table 3:**
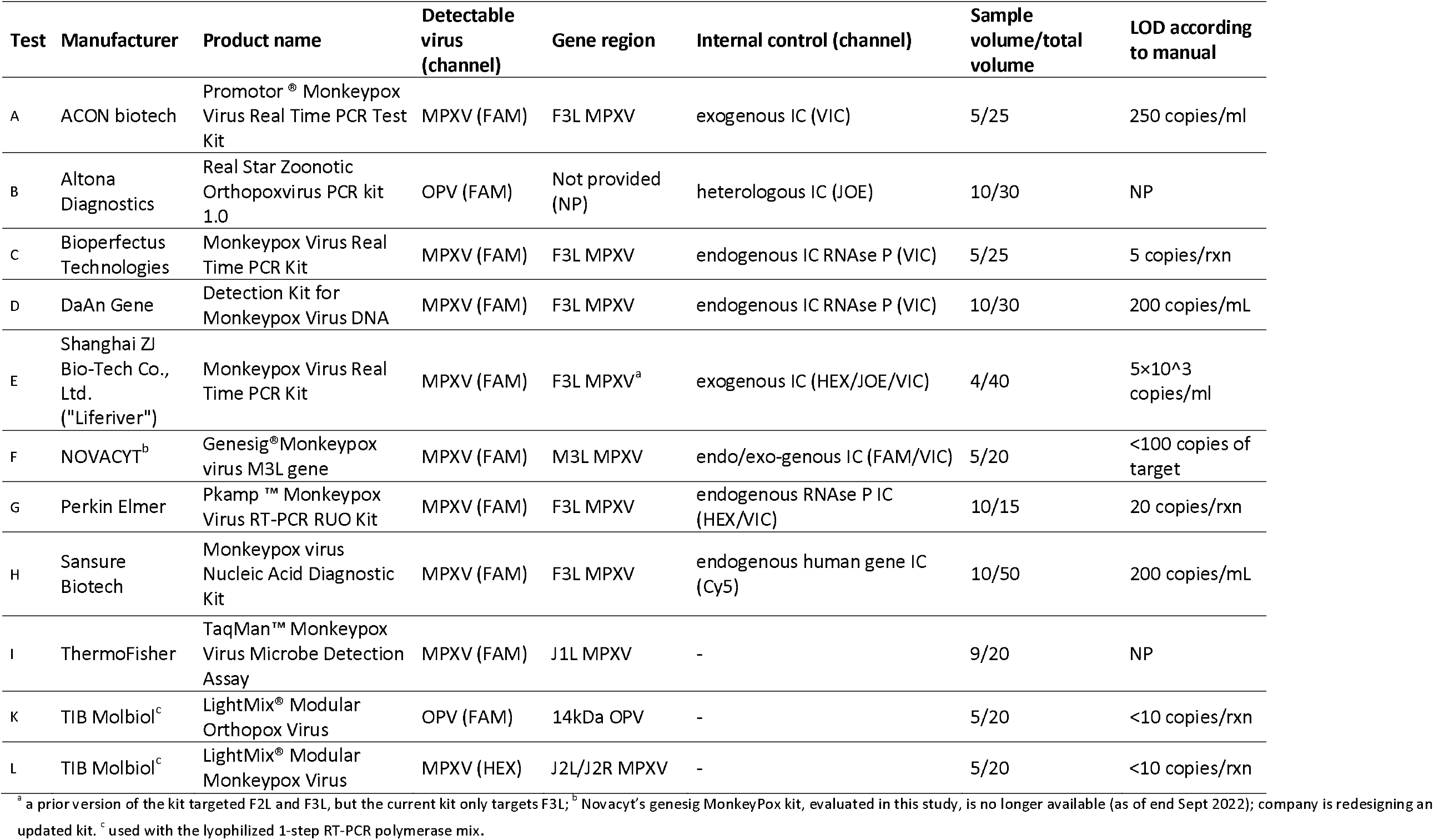
Characteristics of evaluated kits for Monkeypox virus detection

| Test | Manufacturer | Product name | Detectable virus (channel) | Gene region | Internal control (channel) | Sample volume/total volume | LOD according to manual |
| --- | --- | --- | --- | --- | --- | --- | --- |
| A | ACON biotech | Promotor® Monkeypox Virus Real Time PCR Test Kit | MPXV (FAM) | F3L MPXV | exogenous IC (VIC) | 5/25 | 250 copies/ml |
| B | Altona Diagnostics | Real Star Zoonotic Orthopoxvirus PCR kit 1.0 | OPV (FAM) | Not provided (NP) | heterologous IC (JOE) | 10/30 | NP |
| C | Bioperfectus Technologies | Monkeypox Virus Real Time PCR Kit | MPXV (FAM) | F3L MPXV | endogenous IC RNase P (VIC) | 5/25 | 5 copies/rxn |
| D | DaAn Gene | Detection Kit for Monkeypox Virus DNA | MPXV (FAM) | F3L MPXV | endogenous IC RNase P (VIC) | 10/30 | 200 copies/mL |
| E | Shanghai ZJ Bio-Tech Co., Ltd. ("Liferiver") | Monkeypox Virus Real Time PCR Kit | MPXV (FAM) | F3L MPXV<sup>a</sup> | exogenous IC (HEX/JOE/VIC) | 4/40 | 5×10<sup>3</sup> copies/ml |
| F | NOVACYT<sup>b</sup> | Genesig®Monkeypox virus M3L gene | MPXV (FAM) | M3L MPXV | endo/exo-genous IC (FAM/VIC) | 5/20 | <100 copies of target |
| G | Perkin Elmer | Pkamp™ Monkeypox Virus RT-PCR RUO Kit | MPXV (FAM) | F3L MPXV | endogenous RNase P IC (HEX/VIC) | 10/15 | 20 copies/rxn |
| H | Sansure Biotech | Monkeypox virus Nucleic Acid Diagnostic Kit | MPXV (FAM) | F3L MPXV | endogenous human gene IC (Cy5) | 10/50 | 200 copies/mL |
| I | ThermoFisher | TaqMan™ Monkeypox Virus Microbe Detection Assay | MPXV (FAM) | J1L MPXV | - | 9/20 | NP |
| K | TIB Molbiol<sup>c</sup> | LightMix® Modular Orthopox Virus | OPV (FAM) | 14kDa OPV | - | 5/20 | <10 copies/rxn |
| L | TIB Molbiol<sup>c</sup> | LightMix® Modular Monkeypox Virus | MPXV (HEX) | J2L/J2R MPXV | - | 5/20 | <10 copies/rxn |

<sup>a</sup> a prior version of the kit targeted F2L and F3L, but the current kit only targets F3L; <sup>b</sup> Novacyt's genesig MonkeyPox kit, evaluated in this study, is no longer available (as of end Sept 2022); company is redesigning an updated kit. <sup>c</sup> used with the lyophilized 1-step RT-PCR polymerase mix.

The results are summarized in figure 1.

**Figure 1:**
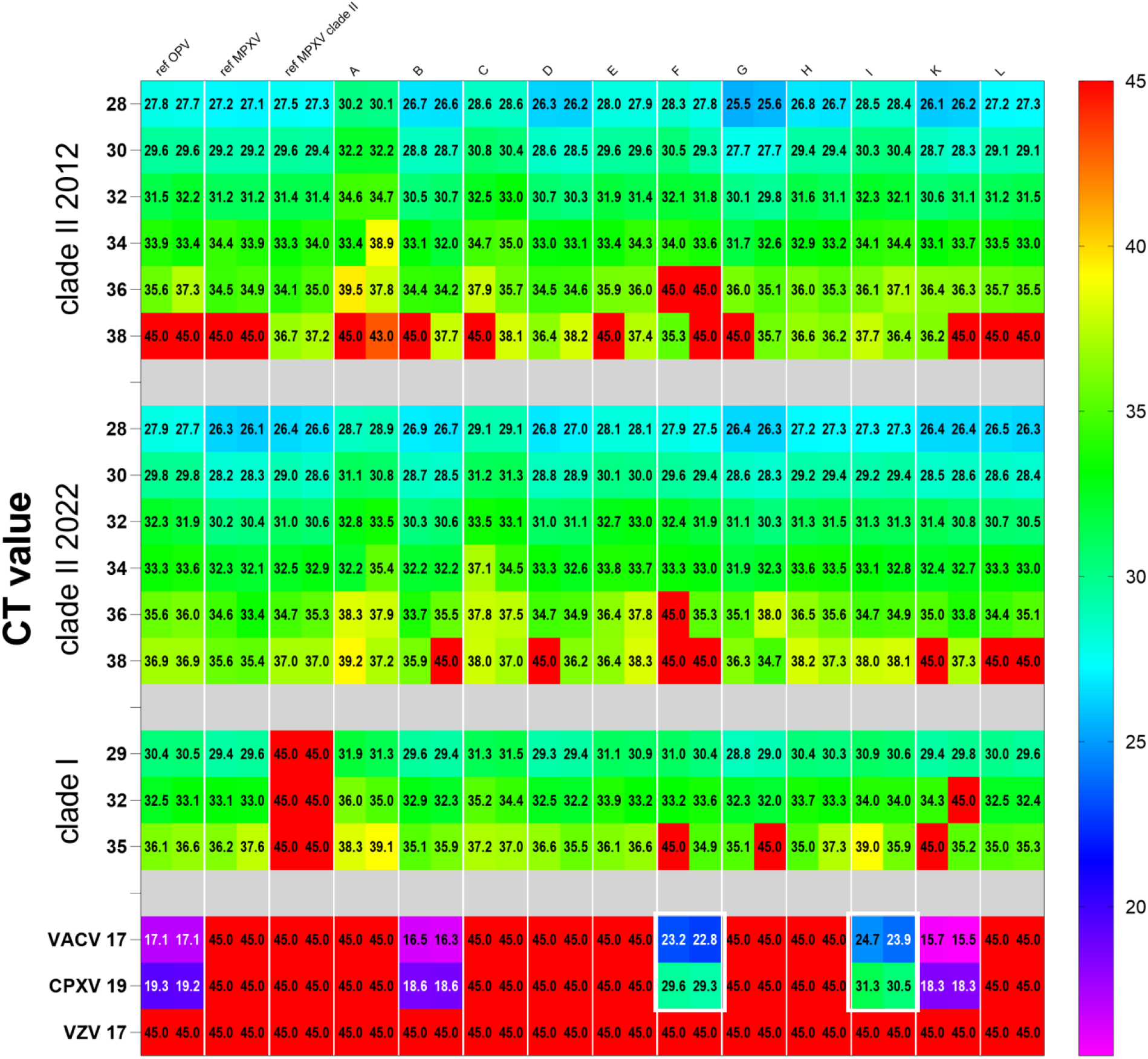
Summary of the CT values obtained for the evaluation panel with the kits A to L. A total of 45 cycles were run, so negative values are indicated in red boxes with CT 45. Bold white boxes highlight cross-reactivity of kits F and I with VACV and CPXV. CT values presented on the left were obtained with the reference PCRs (rpo18 for OPV [5] and in house PCR for VZV).

For the 2012 clade II MPXV isolate, most kits showed CT values in the expected range, indicating good analytical sensitivity down to at least CT 36, reflecting approximately <5 genome equivalents (ge) per reaction (rxn). Only kit F failed to detect this dilution. For the lowest dilution (CT 38, reflecting <1 ge/rxn) two kits and two generic reference PCRs for OPV and MPXV failed to detect both replicates; six kits detected one of two replicates; three kits and the clade II-specific reference PCR detected both replicates, indicating high analytical sensitivity. It should be noted that in this range of DNA concentration, results are prone to higher statistical variation compared to higher MPXV concentrations, and that the dropout of one duplicate may not necessarily reflect poor test performance. Using a 2022 clade II MPXV isolate (IIb), results were similar to those for the 2012 clade II virus, with slightly better detection rates of higher CT samples. Only kit L failed to detect the lowest virus load (CT 38) sample, while the majority of the kits only detected one of the replicates. Clade I MPXV detection was also assessed: kit K failed to detect one duplicate of the CT 32 sample, while three kits failed to detect one duplicate of the lowest concentration (CT 35). Similar CT values were obtained across the different kits for most samples, particularly when considering the varying volumes used per reaction which may contribute to an approximately two-three CT value shift.

Importantly, all controls included in the kits performed as expected. All kits performed within the range specified by the respective manuals. In some manuals, the LOD is given as copies/mL which is not an optimal metric for certain sample types, such as crusts and dry swabs.

Proper sampling of monkeypox lesions generally results in low CT values (high virus loads) [7], therefore the evaluated PCR kits are all likely suitable for diagnosis of MPXV in skin lesions. However, poor sampling may impact the test accuracy; inclusion of endogenous human positive controls in the kits may assist to understand if inadequate sampling occurred in case of a negative result in a suspected patient. Further, sampling at an alternative location may require more sensitive PCR-detection to ensure accurate diagnosis, as the viral kinetics may vary.

We also assessed specificity using VACV, CPXV and VZV DNA. As expected, no kit detected VZV (figure 1). According to the manuals, all kits were designed to be monkeypox virus DNA-specific excluding other OPV, except for kits B and K, which are OPV generic (table 3). Figure 1 confirms that kits B and K also detect VACV and CPXV DNA with CT values similar to the OPV reference PCR [5], whereas the others do not, with two exceptions: kits F and I unexpectedly detected VACV and CPXV DNA, indicating non-specific interactions. We also confirmed the results for kit I on the QuantStudio 5, the thermal cycler specifically recommended by the manufacturer. Both kits F and I resulted in similar CT values, with CT value shifts of approximately 6-7 for VACV and 11-12 for CPXV, indicating better, but still inefficient, binding to VACV DNA than to CPXV DNA. Although the primer and probe sequences were not provided by the manufacturers, kit F targets the M3L gene and kit I targets J1L; a sequence comparison of the M3L monkeypox gene with 92 orthologues of CPXV and 109 VACV showed similarity of 94.6% to 96.8%, and for J1L, comparison of the monkeypox gene with 80 orthologues of CPXV and 99 VACV showed similarity of 83.9% to 97.3%

For further characterization, we plotted the CT values for each DNA sample compared to the calculated number of ge, determined by a plasmid standard curve [8] (supplemental figure 1), and determined the slope, reflecting PCR efficiency (ideally ∼3.32 assuming doubling of PCR product per cycle), and the Y-intercept, indicating the theoretically minimal positive CT value obtained with an assay (table 4).

**Table 4:** Description of standard curves obtained from dilutions of monkeypox virus DNA. Slope and Y-intercept resulting from six, two-fold dilutions of clade II 2012, clade II 2022 and clade I DNA

|  | ref OPV | ref MPXV | ref MPXV<br>clade II | A | B | C | D | E | F | G | H | I | K | L |
| --- | --- | --- | --- | --- | --- | --- | --- | --- | --- | --- | --- | --- | --- | --- |
| **Clade II 2012** |  |  |  |  |  |  |  |  |  |  |  |  |  |  |
| Slope | -3.47 | -3.246 | -3.79 | -3.64 | -3.31 | -3.20 | -3.51 | -3.19 | -2.56 | -3.68 | -3.19 | -2.95 | -3.71 | -3.36 |
| Y intercept | 38.07 | 37.09 | 38.55 | 41.18 | 36.56 | 38.40 | 36.88 | 37.47 | 36.17 | 36.65 | 36.83 | 37.51 | 37.52 | 37.32 |
| **Clade II 2022** |  |  |  |  |  |  |  |  |  |  |  |  |  |  |
| Slope | -2.999 | -3.109 | -3.165 | -2.558 | -3.295 | -3.455 | -3.31 | -3.34 | -3.82 | -3.67 | -3.18 | -2.95 | -3.70 | -3.36 |
| Y intercept | 37.08 | 36.41 | 37.87 | 36.17 | 36.49 | 37.66 | 37.17 | 36.61 | 39.38 | 36.65 | 36.83 | 37.51 | 37.52 | 37.32 |
| **Clade I** |  |  |  |  |  |  |  |  |  |  |  |  |  |  |
| Slope | -3.18 | -3.99 |  | -3.83 | -3.26 | -3.08 | -3.63 | -2.91 | -2.37 | -3.35 | -3.13 | -3.59 | -3.21 | -2.90 |
| Y intercept | 38.9 | 40.3 | 40 | 42.12 | 38.37 | 39.94 | 39.1 | 38.85 | 37.33 | 38.07 | 38.95 | 40.5 | 38.67 | 37.65 |

Supplemental Figure 1: Standard curves for the dilution series of MPXV clade II from 2012 (A), clade II from 2022 (B) and clade I (C) viruses. Copy number expressed in genome equivalents per reaction (ge/rxn) are plotted vs the CT value.

Small variations in CT values per sample were observed, however, in general all kits resulted in comparable standard curves, particularly for both clade II viruses. An exact quantification of virus DNA in lesions and other tissue samples is hampered by the lack of a reference standard, but quantification in primary poxvirus diagnostics is not of great relevance.

## Conclusion

The eleven kits evaluated show comparable and high sensitivity to detect clade I and clade II monkeypox virus DNA, and therefore were found to be suitable to identify a range of clinically-relevant viral loads of monkeypox virus DNA for diagnosis using properly-sampled skin lesions. Analytical sensitivity of the kits was generally high detecting down to less than approximately 5 ge/rxn (CT 36) and the limited specificity assessment showed most assays to be specific for MPXV or OPV, as per their intended design. It should be noted that the included kits and the many others coming to market are currently intended for research use only; the generation and dissemination of data assessing clinical performance continues to be needed to ensure increased adoption of accurate kits that enable communities globally who are most affected to access diagnosis of monkeypox virus.

## Data Availability

All data produced in the present study are available upon reasonable request to the authors

## Author contributions

Janine Michel, Jilian A. Sacks, Lars Schaade and Andreas Nitsche conceptualized the study, the evaluation panel, selected the viruses, analysed the data and wrote the manuscript. Daniel Bourquain propagated and quantified the monkeypox virus strains. Angelina Targosz and Thomas Rinner prepared the evaluation panel und performed the PCR experiments. Annika Brinkmann sequenced the genomes of the used virus variants and analysed the primer and probe binding to Orthopoxviruses. All authors read and approved the final version of the manuscript.

## Acknowledgement

The authors thank WHO (Laboratory Team, Emerging Diseases and Zoonoses Unit of the Health Emergencies Programme, led by Dr. Mark Perkins and including Dr. Lorenzo Subissi) for providing the kits evaluated and for technical input around study design and result interpretation. The authors are grateful to Ursula Erikli for copy-editing.

## References

1. Carroll DS, Emerson GL, Li Y, Sammons S, Olson V, Frace M, et al. Chasing Jenner’s vaccine: Revisiting Cowpox virus classification. PLoS One. 2011;6(8).

2. Reed KD, Melski JW, Graham MB, Regnery RL, Sotir MJ, Wegner M V, et al. The Detection of Monkeypox in Humans in the Western Hemisphere. N Engl J Med [Internet]. 2004;350(4):342–50. Available from: https://doi.org/10.1056/nejmoa032299

3. Monkeypox: WHO declares a public health emergency of international concern (PHEIC) [Internet]. 2022. Available from: https://www.who.int/europe/news/item/23-07-2022-who-director-general-declares-the-ongoing-monkeypox-outbreak-a-public-health-event-of-international-concern

4. Li Y, Zhao H, Wilkins K, Hughes C, Damon IK. Real-time PCR assays for the specific detection of monkeypox virus West African and Congo Basin strain DNA. J Virol Methods [Internet]. 2010/07/17. 2010 Oct;169(1):223–7. Available from: https://www.ncbi.nlm.nih.gov/pubmed/20643162

5. Schroeder K, Nitsche A. Multicolour, multiplex real-time PCR assay for the detection of human-pathogenic poxviruses. Mol Cell Probes [Internet]. 2009/10/29. 2010 Apr;24(2):110–3. Available from: https://www.ncbi.nlm.nih.gov/pubmed/19879351

6. Kurth A, Nitsche A. Detection of human-pathogenic poxviruses. Methods Mol Biol. 2011;665:257–78.

7. Tarín-Vicente EJ, Alemany A, Agud-Dios M, Ubals M, Suñer C, Antón A, et al. Clinical presentation and virological assessment of confirmed human monkeypox virus cases in Spain: a prospective observational cohort study. Lancet (London, England). 2022 Aug;400(10353):661–9.

8. Nitsche A, Ellerbrok H, Pauli G. Detection of Orthopoxvirus DNA by Real-Time PCR and Identification of Variola Virus DNA by Melting Analysis. J Clin Microbiol. 2004;42(3):1207–13.

