## Supplementary figures and images for "Evaluation of eleven commercially available PCR kits for the detection of Monkeypox virus DNA"

### Supplemental Figure 1

#### Supplemental figure 1

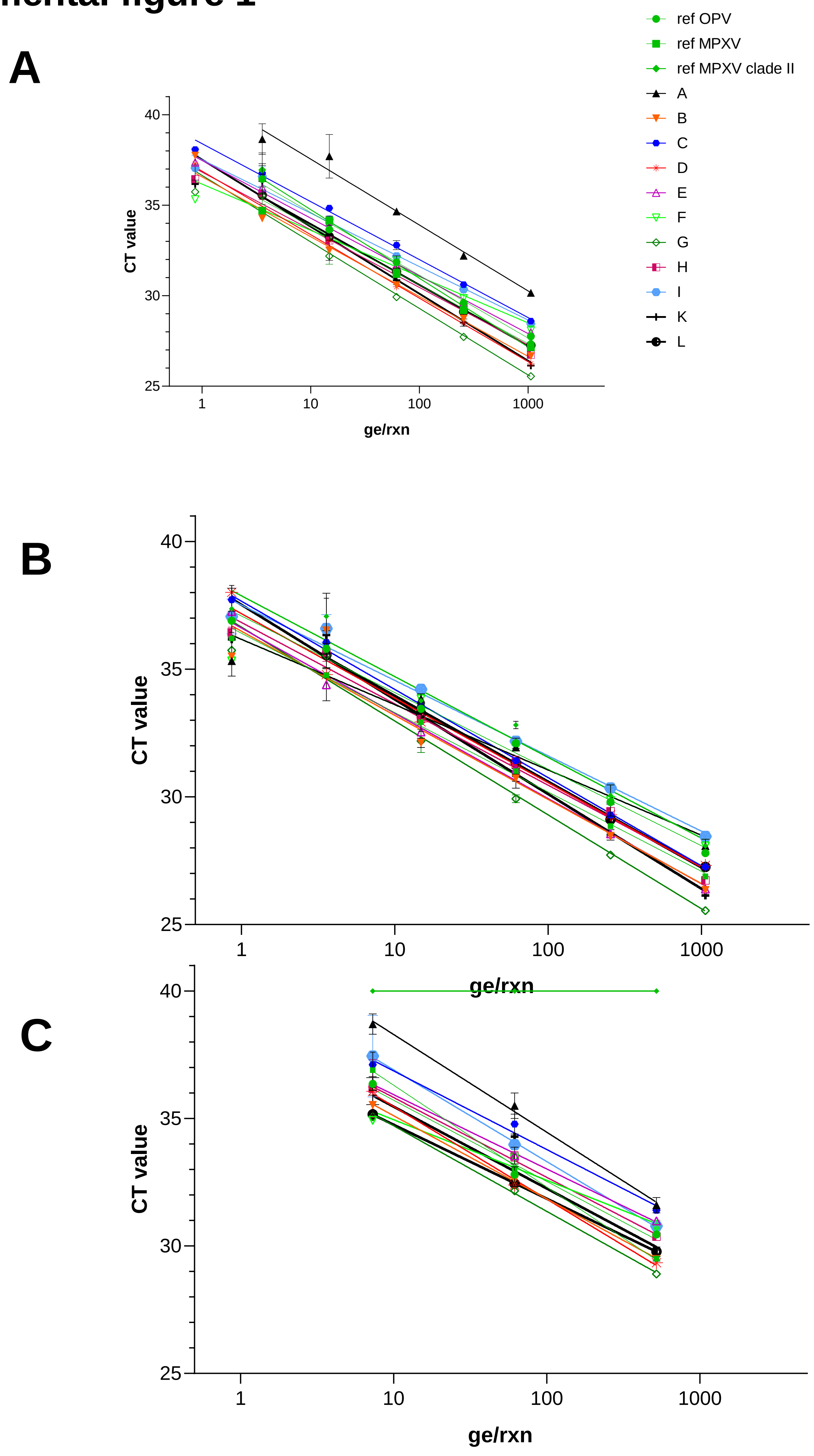
